# BODY SURFACE GASTRIC MAPPING DELINEATES SPECIFIC PATIENT PHENOTYPES IN ADOLESCENTS WITH FUNCTIONAL DYSPEPSIA AND GASTROPARESIS

**DOI:** 10.1101/2024.05.13.24307307

**Authors:** Gayl Humphrey, Celia Keane, Gabriel Schamberg, Alain Benitez, Stefan Calder, Binghong Xu, Christian Sadaka, Christopher N. Andrews, Greg O’Grady, Armen Gharibans, Hayat Mousa

## Abstract

**Importance:** Persistent upper gastroduodenal symptoms, such as nausea, vomiting, bloating, and abdominal pain, are widespread among pediatric patients. Multiple overlapping symptoms complicate the diagnostic process, necessitating the development of novel gastric function tests with actionable biomarkers. Body Surface Gastric Mapping (BSGM) has emerged as a promising diagnostic tool for gastroduodenal disorders, and this is the first detailed evaluation in adolescents.

**Objective:** This study aimed to assess the utility of BSGM in delineating specific patient phenotypes among adolescents with functional dyspepsia (FD) and gastroparesis in order to guide clinical decision-making.

**Design:** A prospective cross-sectional study recruited adolescents aged 12 to 21 between 2022 and 2024.

**Setting:** Controls were recruited from New Zealand (controls) and Patients from the Children’s Hospital of Philadelphia, USA.

**Participants:** Prospectively recruited participants included controls without gastroduodenal symptoms or motility-related medication usage and patients diagnosed with either gastroparesis (delayed gastric emptying test (GET)) or FD according to ROME IV criteria and a normal GET.

**Procedures:** BSGM was performed using a standardized protocol, including simultaneous symptom reporting and the completion of validated symptom, psychometric and physical health questionnaires.

**Main Outcome:** The primary outcome was to evaluate if BSGM could delineate specific patient phenotypes and provide clinically meaningful distinctions between gastroparesis and FD diagnoses, utilizing BSGM spectral outcome data.

**Results:** Fifty-six subjects were recruited (31 controls, 25 patients); median age 16; 96% of patients were female. Control data showed that adult reference intervals provided an acceptable interpretation framework. Patients with FD (n=10) and gastroparesis (n=15) had common symptoms, mental health, quality of life and functional disability (all p>0.05). Three distinct BSGM phenotypes were identified: *BSGM Normal* (n=10), *BSGM Delay* (n=8), and *Low Stability/Low Amplitude* (n=7), having spectral differences in BMI-Adjusted Amplitude 34.6 vs 39.1 vs 19.9 (*p*=.01) and Gastric Alimetry Rhythm Index: 0.45 vs 0.45 vs 0.19 (*p*=.003).

BSGM phenotypes demonstrated differences in symptoms (nausea *p*=0.04), physical health (*p*=.04) and psychometrics (anxiety *p*=.03).

**Conclusion and Relevance:** Adolescent patients with FD and gastroparesis have overlapping clinical profiles, making individualized treatment challenging. Conversely, employing BSGM to categorize patients into distinct phenotypes revealed clinically relevant differences, offering potential avenues for individualized therapeutic pathways.

## Introduction

Persistent upper gastroduodenal symptoms such as nausea, vomiting, bloating and abdominal pain are prevalent in the pediatric population,^1^ impacting quality of life and leading to frequent healthcare presentations.^2, 3^ The Rome IV pediatric criteria provide a diagnostic framework to support a positive diagnostic approach;^4^ however, overlapping symptoms and diagnostic criteria continue to pose challenges to personalized treatment.^5^

Per Rome IV, FD is subclassified into postprandial distress syndrome (PDS) and epigastric pain syndrome (EPS), which is not explicitly related to food intake.^4^ However, approximately 35% of FD patients experience both PDS and EPS.^6^ Patients with gastroparesis also commonly report epigastric pain and postprandial distress, in addition to nausea and vomiting, while demonstrating delayed gastric emptying.^7^ However, up to 25% of patients with FD also show delayed emptying, underscoring an overlapping pathophysiology.^8^ Gastric emptying as a diagnostic standard has also been challenged due to questions regarding reproducibility^9^ and symptom correlations.^10, 11^ New gastric tests providing actionable biomarkers are needed to beter discriminate disease pathophysiology to inform care.

Body Surface Gastric Mapping (BSGM) (Gastric Alimetry™, Alimetry, New Zealand) is a new diagnostic test involving high-resolution electrophysiology coupled with simultaneous symptom profiling. Several studies applying BSGM in adults have demonstrated the capability to phenotype specific disease subgroups in gastroduodenal disorders.^12, 13^ A recent study also demonstrated that using BSGM phenotypes in clinical practice aided decision-making and reduced healthcare costs.^14^ In addition, in a large head-to-head study, BSGM demonstrated a higher yield for gastric motility abnormalities than gastric emptying testing (GET) alone, with improved correlations to symptoms and psychometrics.^9^ Feasibility studies using BSGM in pediatrics have been presented in abstract form, suggesting that BSGM is acceptable, feasible, and safe for children as young as five.^15, 16^ This study is the first detailed report on using BSGM with a pediatric population, focusing on adolescents. The aims were to define whether BSGM can delineate specific patient subgroups within FD and gastroparesis and to compare symptoms, health psychology, and quality of life across these subgroups.

## Methods

This was a pragmatic cross-sectional study, with a sample size estimated at a minimum of 50 subjects from recent feasibility studies in pediatric populations.^15, 16^ The study is reported in accordance with the STROBE guidelines.^17^

### Population

Adolescent participants were recruited from Auckland, New Zealand (controls) and the Children’s Hospital of Philadelphia, USA (patients). Ethics was approved by the relevant institutional review boards, and all participants provided informed consent. Healthy controls were recruited through local advertising and patient participants through clinical referrals.

Eligibility criteria were that participants were aged between 12-21 years, had a body mass index (BMI) <35kg/m^2^ and had no history of gastric surgery other than percutaneous enteric gastrostomy tube insertion (allowed for patients). Patient eligibility also included FD (confirmed by Rome IV and a normal GET) or a clinical diagnosis of gastroparesis (confirmed by a delayed GET). All GET were undertaken within 24 months of the BSGM test.

### Procedures

Participant demographics, anthropometric measures, and clinical data, including GET outcomes, were obtained. A delayed GET confirmed gastroparesis and Rome IV criteria, and a normal GET confirmed FD.^18, 19^ All participants completed age-appropriate validated questionnaires for gastroduodenal symptoms, functional disability, mental wellbeing and quality of life on the day of the test.

Figure 1 presents the standardized BSGM protocol using the Gastric Alimetry system (details previously published),^20^ screenshots of the symptom-logging app and an example of the array on an adolescent abdomen. The standard test protocol involved gentle skin exfoliation with an electrolyte gel, a 30-minute fasting baseline, a standardized meal (68 g Clif® bar; 250kCal with 45g carbohydrate, 9g protein, 5g fat, 4g fibre) and 200mls of water, consumed over 10 minutes, followed by a 4-hr postprandial recording. Simultaneously, all participants reported their symptoms on the Gastric Alimetry App using an 11-point Likert scale of 0=’none’ to 10=’worst symptom imaginable’ and reported episodic symptoms (belching, reflux, vomiting) at a minimum of 15-minute intervals. The total symptom burden score and individual symptom scores were calculated and averaged.^21^

**Figure 1.**
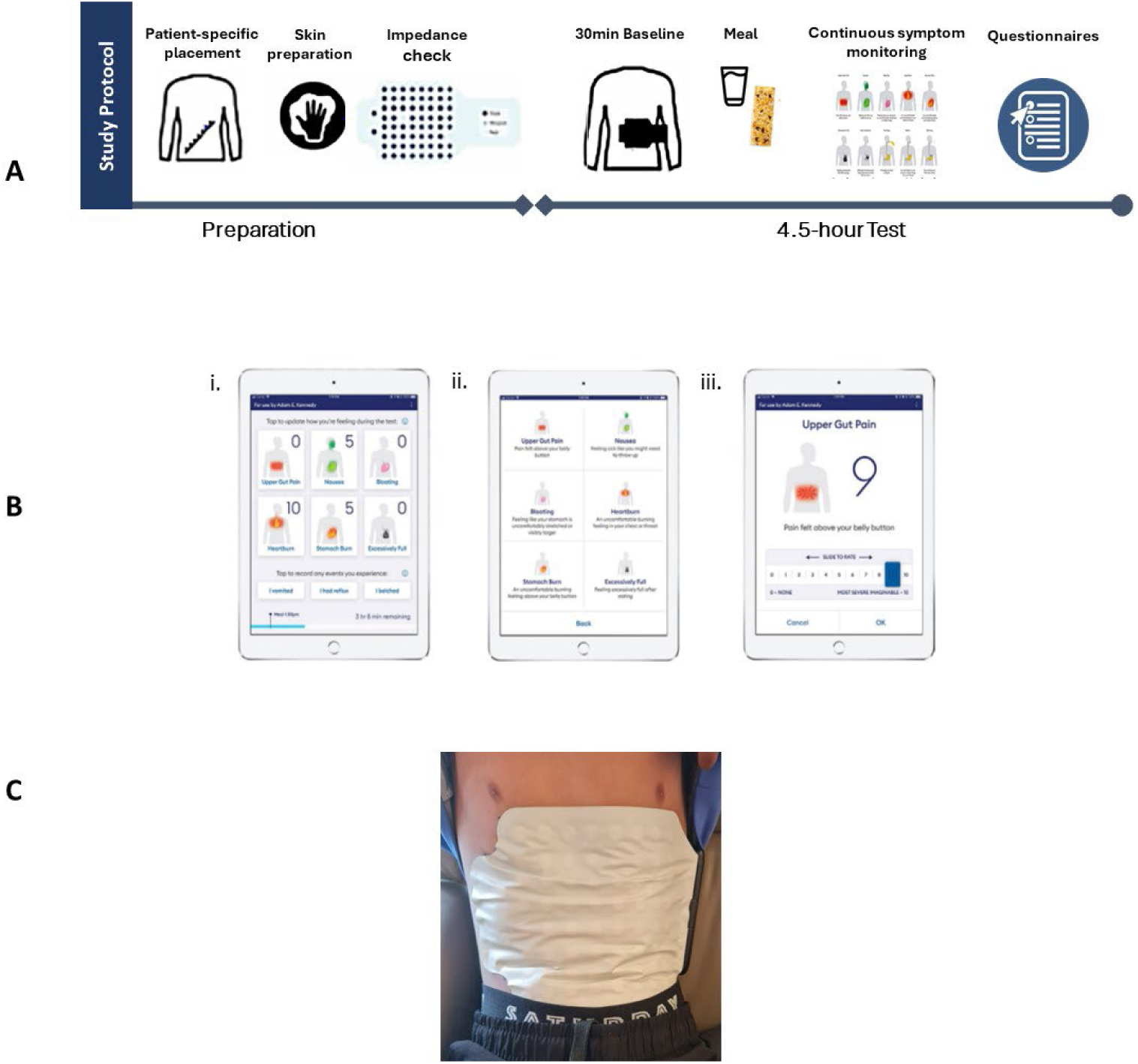
A. Standardized BSGM study protocol, B screenshots of the Alimetry Symptom-Logging App; i. the symptom reporting dashboard, ii. the symptoms explained, and iii. the upper gut pain symptom displayed, and C. the electrode dense hydrogel array placed on a male aged 13 years.

BSGM spectral metrics are reported as Principal Gastric Frequency (PGF), BMI-Adjusted Amplitude, Gastric Alimetry Rhythm Index (GA-RI) and fed-fasted Amplitude Ratio (ff-AR) (for technical details refer to ^20, 22^). In the absence of validated pediatric normative reference intervals, abnormalities were defined using adult reference intervals and visual assessment of the spectral plots.^12, 20^ Additionally, the adolescent control data generated in this study were compared to adult normative ranges to ensure that these provided an acceptable provisional interpretative framework. Patients were then phenotyped a priori according to abnormalities in the BSGM metrics, symptoms, and meal response pattern based on previous approaches used in adults.^13^

### Analysis

Statistical analyses were performed using SPSS version 29.0 (IBM Corporation, Armonk, NY). Variables were summarised with descriptive statistics, normality was assessed, and data expressed as mean with standard deviation (SD) or median and interquartile range (IQR).

Statistical comparisons were performed between study groups using Student’s t-test, Pearson’s chi-squared tests, Mann-Whitney U, Kruskal-Wallis, or Fisher’s exact tests as appropriate. Pearson’s correlation coefficient and Spearman’s correlation were also applied as appropriate to data type.

## Results

56 subjects comprising 31 healthy controls and 25 patients were recruited; median age 16 years (range 12-20); 80% were female and 74% Caucasian (eTable 1). Mean BMI was not different between the groups. Summary patient and control data for total symptom burden score, quality of life and mental wellbeing are presented in eTable 2 and are similar to that reported in other international studies.

The previously developed adult reference intervals^20, 22^ were compared to 31 healthy control adolescents aged 12-18 years to verify the acceptability of their use in adolescents. The main difference was that pediatric controls showed a modestly lower median GA-RI than adults (0.35 (0.22-0.43) versus 0.50 (0.39-0.64), *p*<0.001; eTable3). However, as this study’s aim is to detect group-level distinctions, the overall similarity between pediatric and adult controls provided confidence that applying the adult reference intervals could discern phenotypes and meaningful group-level differences within the pediatric patient cohort.

### Gastroparesis vs Functional Dyspepsia

All 25 subjects underwent GET, with 15 having gastroparesis and 10 having normal GET (defined as FD, PDS n=5, or EPS n=5 as per Rome IV). There were no significant differences in total symptom burden, individual symptoms during the test, clinical symptoms, quality of life, functional disability, or mental wellbeing between patients with gastroparesis or FD. Likewise, BSGM metrics were not significantly different between gastroparesis or FD patient groups (see Table 1). We also found no significant differences when subgrouping FD into EPS (PGF (mean 2.84 ± .17 vs 3.01 ± 1.6; *p*=0.7); BMI-Adjusted Amplitude (27.3 ±6.9 vs 31.2 ± 9.2; p=0.4); GA-RI 0.30 ± .18 vs 0.38 ± .28; *p*=0.3), or ff-AR (1.5 ± .37 v 1.4 ±.94; *p*=0.6).

**Table 1.** BSGM within-test total symptom burden and individual symptom scores; clinical symptoms, Quality of Life, Functional Disability, and Mental Wellbeing questionnaire outcomes and BSGM spectral metric outcomes for Functional Dyspepsia and Gastroparesis Patients.

| VARIABLES (median (IQR)) |  | Functional Dyspepsia<br>n=10 | Gastroparesis<br>n=15 | P value |
| --- | --- | --- | --- | --- |
| **Within-Test Symptoms** |  |  |  |  |
| Total Symptom Burden Score |  | 27.50 (15.7-33.2) | 22.8 (5.0-39.4) | .95 |
| Individual Symptoms | Early Satiety | 5 (1.0-7.0) | 4.1 (3-6) | .48 |
|  | Excessively Full | 2.7 (2.4-6.5) | 1.3 (.1-5.8) | .80 |
|  | Bloating | 3.5 (1.8-4) | 1.8 (.5-4.9) | .55 |
|  | Heartburn | 0.5 (0.0-2.0) | 0.5 (0.0-4.1) | .34 |
|  | Nausea | 3.6 (2.3-5.9) | 4.5 (.3-7.1) | .79 |
|  | Upper Gut Pain | 4.3 (2.0-5.2) | 2.2 (0.0-6.7) | .88 |
|  | Stomach Burn | 0.3 (0.0-5.0) | 1.1 (0.0-4.6) | .39 |
| **Patient Completed Questionnaires** |  |  |  |  |
| Gastrointestinal Symptoms Module | ↓ | 54.2 (33.1-68.5) | 47.3 (38.5-71.2) | .57 |
| Nausea Severity Scale | ↑ | 27.0 (19.7-29.2) | 30.0 (20.0-32.0) | .71 |
| Abdominal Pain Index | ↑ | 25.0 (12.5-33.0) | 25.1 (15.0-30.0) | .34 |
| Functional Disability Index | ↑ | 23.5 (11.7-35.0) | 27.0 (9.0-34.0) | .88 |
| Quality of Life | ↓ | 60.0 (45.0-74.5) | 53.0 (47.0-63.0) | .16 |
| Anxiety | ↑ | 58.3 (43.3-61.8) | 54.8 (47.2-61.8) | .58 |
| Depression | ↑ | 50.7 (43.5-65.1) | 49.8 (37.7-62.6) | .48 |
| **BSGM Spectral Data** |  |  |  |  |
| Principal Gastric Frequency (cpm)<sup>a</sup> |  | 2.90 (2.84-2.99) | 3.05 (2.95-3.13) | .21 |
| BMI-Adj Amplitude (μV)<sup>b</sup> |  | 26.6 (24.5-31.8) | 40.4 (27.9-48.2) | .06 |
| Gastric Alimetry Rhythm Index |  | .31 (.19-.36) | .47 (.32-.63) | .13 |
| Fed to Fasted Amplitude Ratio |  | 1.73 (1.49-1.86) | 1.81 (1.31-2.35) | .98 |
| ↑↓ : Direction of severity for each questionnaire: <sup>a</sup>cpm: cycles per minute; <sup>b</sup>μV: microvolts |  |  |  |  |

Figure 2 presents the averaged spectral and symptom burden plots derived from the Gastric Alimetry test for controls and patients with FD and gastroparesis.

**Figure 2.**
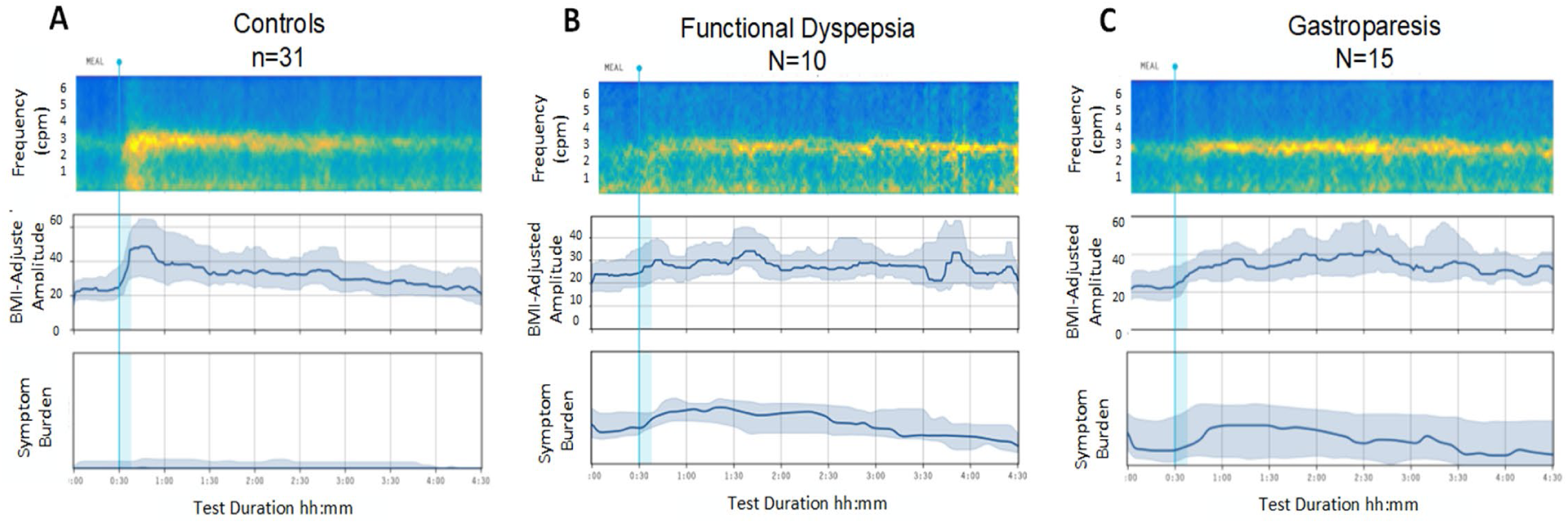
Average spectrogram, median BMI-Adjusted Amplitude (IQR shaded), and mean Symptom Burden (SD shaded) for (**A**) Healthy Controls and (**B**) FD and (**C**) Gastroparesis patients.

Gastroparesis and FD subjects were, therefore, clinically indistinguishable across symptom severity, functional disability, psychometric profiles, and BSGM spectral metrics.

### BSGM: Phenotype Subgroups Analysis

The 25 patients were defined and categorized a priori into three distinct phenotypes according to their spectral metrics and symptom profiles, regardless of FD or gastroparesis status, as follows:

- *BSGM Normal* (n=10): indistinguishable spectral metrics to healthy controls
- *BSGM Delayed* (n= 8): delay in onset of gastric activity postprandially defined by meal response ratio <1 (ratio of amplitude in the first 2 hours postprandially to the last 2 hours).
- *BSGM Low Stability/Low Amplitude* (n=7): have a GA-RI <0.25 and/or BMI-Adjusted amplitude <22µv

There were no differences in age, BMI, or test quality measures between phenotypes (eTable 4). BSGM metrics, symptoms, and questionnaire scores are provided in Table 2. The phenotyped BSGM groups demonstrated significant differences in psychology and physical health metrics, including anxiety scores, which were worse for *the Low Stability / Low Amplitude* phenotype (59.5 (51.8-61.8) than *BSGM Delayed* (57.4 (47.5-59.5) and *BSGM Normal* (43.5 (36.8-47.2), *p*=0.03). A similar patern emerged with Functional Disability scores, with *Low Stability / Low Amplitude* phenotype reporting a higher impact on functional ability (21.5 (8.4-26.5) than *BSGM Delayed* 18.5 (2.3-36.8) and *BSGM Normal* (12.5 (3.0-24), *p*=0.04).

**Table 2.** Spectral data, total symptom burden, individual symptom scores, clinical symptoms, Quality of Life, Functional Disability, and Mental Wellbeing Questionnaire Outcomes by Phyotypes, BSGM Normal, BSGM Delayed and Low Stability/Low Amplitude.

| VARIABLES<br>(median (IQR)) | PHENOTYPES |  |  | BSGM<br>Normal v<br>Delayed<br>v LS/LA | BSGM<br>Normal<br>v BSGM<br>Delayed | BSGM<br>Normal<br>v LS/LA | BSGM<br>Delayed<br>v LS/LA |
| --- | --- | --- | --- | --- | --- | --- | --- |
|  | BSGM<br>Normal<br>N=10 | BSGM Delayed<br>N=8 | LS/LA<sup>a</sup><br>N=7 |  |  |  |  |
|  |  |  |  | P value | P value | P value | P value |
| **BSGM METRICS** |  |  |  |  |  |  |  |
| PGF (cpm)<sup>b</sup> | 3.00<br>(2.86-3.05) | 2.96<br>(2.89-3.17) | 2.84<br>(2.62-3.05) | .25 | .89 | .23 | .21 |
| BMI-Adj<br>Amplitude (μV)<sup>c</sup> | 34.6<br>(30.0-45.7) | 39.05<br>(26.7-49.4) | 19.8<br>(17.1-29.5) | .01 | .88 | .06 | .006 |
| GA-RI<sup>d</sup> | 0.45<br>(.31-.52) | 0.45<br>(.33-.57) | 0.19<br>(.16-.19) | .003 | .63 | <.001 | <.001 |
| ff-AR<sup>e</sup> | 1.65<br>(1.19-3.36) | 1.84<br>(1.21-2.12) | 1.56<br>(1.21-2.12) | .29 | .94 | .12 | .16 |
| Meal Response<br>Ratio | 1.37<br>(1.11-1.44) | 0.84<br>(0.81-0.95) | 1.06<br>(1.01-1.34) | .004 | .01 | .48 | .048 |
| **SYMPTOMS** |  |  |  |  |  |  |  |
| Total Symptom<br>Burden Score | 18.35<br>(5.7-27.9) | 25.10<br>(10.5-27.5) | 33.2<br>(17.1-38.7) | .08 | .76 | .055 | .15 |
| Early Satiety | 4.5<br>(2.5-6.0) | 3.0<br>(.25-5.7) | 6.0<br>(0.0-8.0) | .72 | .52 | .35 | .71 |
| Excessively Full | 0.9<br>(.07-3.2) | 2.55<br>(.25-5.7) | 3.30<br>(1.3-7.5) | .49 | .48 | .09 | .57 |
| Bloating | 1.9<br>(.37-2.8) | 1.45<br>(0.0-3.4) | 4.00<br>(.4-7.1) | .43 | .60 | .04 | .09 |
| Heartburn | 0.3<br>(.0-2.7) | 0.4<br>(.02-2.7) | 0.3<br>(0.0-4.1) | .60 | .65 | .64 | .90 |
| Nausea | 1.4<br>(.07-4.1) | 3.5<br>(.15-6.9) | 5.5<br>(4.8-9.1) | .04 | .38 | .04 | .26 |
| Upper Gut Pain | 1.35<br>(0.0-5.2) | 2.05<br>(0.0-6.2) | 4.9<br>(1.6-5.7) | .06 | .11 | .03 | .16 |
| Stomach Burn | 0.0<br>(0.0-3.7) | 0.75<br>(0.0-5.3) | 0.0<br>(.0-2.5) | .69 | .3 | .92 | .38 |
| **SYMPTOMS, QUALITY OF LIFE, FUNCTIONAL DISABILITY AND PSYCHOMETRIC QUESTIONNAIRES** |  |  |  |  |  |  |  |
| Gastrointestinal<br>Symptoms | 66.22<br>(40.88-73.82) | 46.28<br>(28.72-47.30) | 54.22<br>(38.51-66.22) | .33 | .55 | .88 | .71 |
| Nausea Severity<br>Scale | 15.0<br>(2.2-25.7) | 25.5<br>(14.3-30.5) | 29.5<br>(20.0-32.0) | .57 | .44 | .32 | .81 |
| Abdominal Pain<br>Index | 20.0<br>(11.3-27.3) | 21.0<br>(6.25-29.5) | 28.2<br>(3.75-33.0) | .06 | .75 | .06 | .50 |
| Functional<br>Disability Index | 12.5<br>(3.0-24.0) | 18.5<br>(2.3-36.8) | 21.5<br>(8.7-26.5) | .04 | .087 | .042 | .73 |
| Quality of Life | 63.5<br>(45.3-85.7) | 59.5<br>(50.7-75.5) | 57.5<br>(46.3-76.3) | .06 | .09 | .06 | .72 |
| Anxiety | 43.5<br>(36.8-47.2) | 57.4<br>(47.5-59.5) | 59.5<br>(51.8-61.8) | .03 | .09 | .03 | .5 |
| Depression | 43.4<br>(37.7-56.7) | 52.3<br>(40.7-63.3) | 50.7<br>(43.9-65.6) | .56 | .10 | .33 | .88 |

<sup>a</sup>LS/LA: Low stability/Low amplitude; <sup>b</sup>PGF: Principal Gastric Frequency; cpm: cycles per minute; <sup>c</sup>μV: microvolts; <sup>d</sup>GA-RI: Gastric Alimetry Rhythm Index; <sup>e</sup>ff-AR: fed to fasted Amplitude Ratio.

This patern was repeated in abdominal pain severity index scores (20.0 (11.3-27.3) vs 21.0 (6.25-29.5) vs 28.2 (3.75-33.0), *p*=.06), and quality of life (63.5 (45.3-85.7) vs 59.5 (50.7-75.5) vs 57.5 (46.3-76.3), *p*=0.06), although not reaching statistical significance.

Within-test symptom comparisons between the phenotyped groups found that nausea was highest in the *Low Stability/Low Amplitude* group (5.5 vs *BSGM Delayed* 3.5 vs *BSGM Normal* 1.4, *p*=0.04). The total symptom burden score was also higher in the *Low Stability/Low Amplitude* group (33.2 (17.1-38.7) vs *BSGM Delayed* 25.1 (10.5-27.5) and *BSGM Normal* 18.3 (5-7-27.9), although not reaching statistical significance: *p*=0.08). Pairwise analyses showed higher symptom severity in the *Low Stability/Low Amplitude* group compared to *BSGM Normal* for nausea (5.5 (4.8-9.1) vs 1.4 (.07-4.1), *p*=0.04), upper gut pain (4.9 (1.6-5.7) vs 1.35 (0.0-3.7), *p*=0.03) and bloating (4.0 (.4-7.1) vs 1.9 (.37-2.8), *p*=0.04). The remaining symptoms did not differ between groups; however, there are visible differences in the symptom curves for each symptom by phenotype (eFigure 1).

Figure 3 A-C displays the average spectral and symptom data for each BSGM phenotype. *BSGM Normal* (Figure 3A) showed a high symptom burden that was present pre-prandially and continued post-prandially, being moderately meal-responsive and with no correlation between the gastric amplitude and symptom curves (Spearman’s correlation r=0.11 *p*=0.7 (95%CI -0.53-0.7)). *BSGM Delayed* (Figure 3B) showed an increase in symptoms postprandially, which decreased as gastric amplitude increased (Spearman’s correlation r= -0.26, *p*=.06, 95%CI -0.18–0.54). The *Low Stability/Low Amplitude* phenotype (Figure 3C) showed a relatively high symptom burden pre-prandially, which remained continuous throughout the test (total symptom burden score 33.2) and with symptom curves uncorrelated with gastric amplitude (Spearman’s correlation r=0.21, *p=*0.65 (95%CI -0.78-0.55).

**Figure 3.**
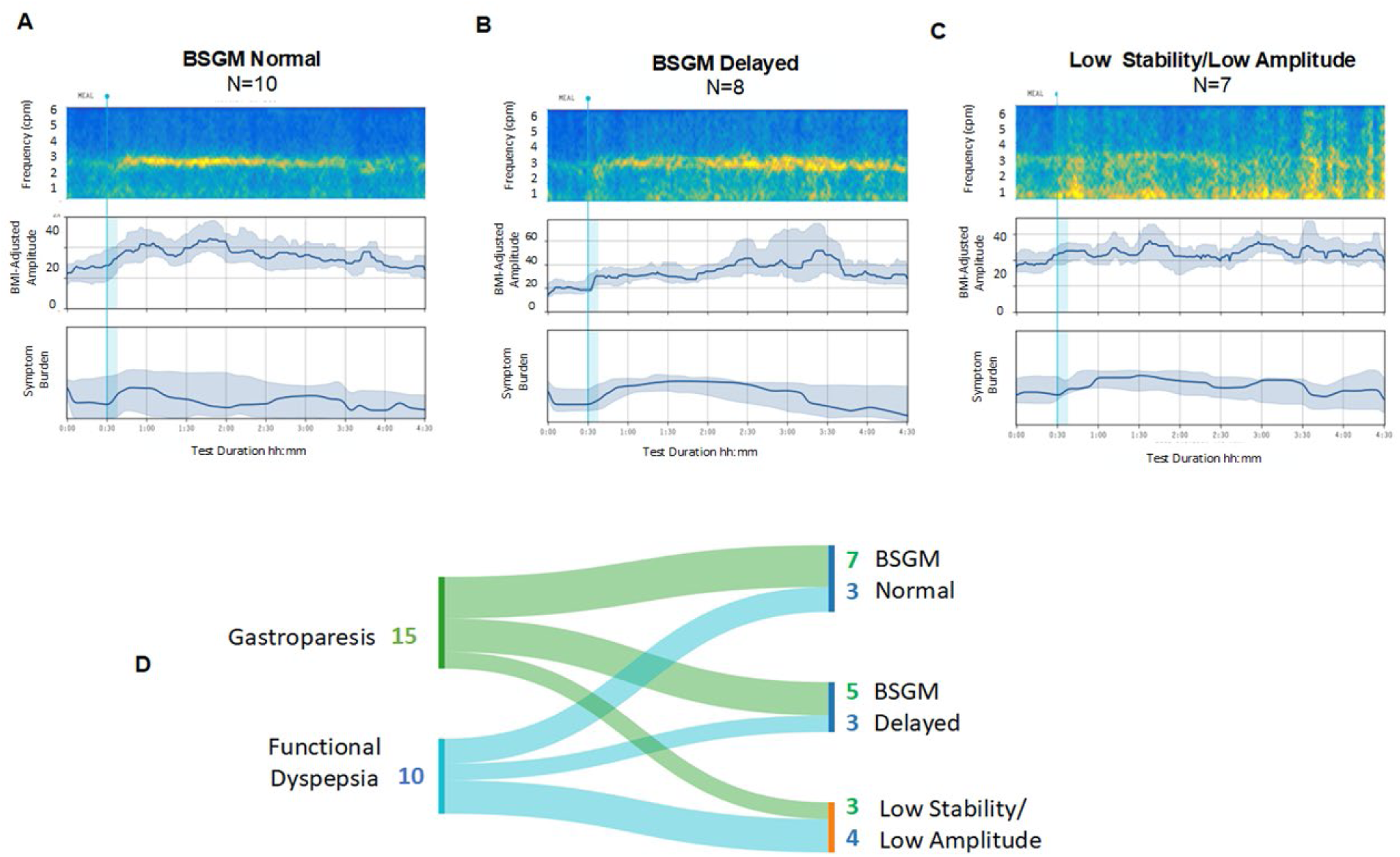
Figures **A-C** presents the average spectrograms, median BMI-adjusted amplitude curves (IQR shaded) indicating meal responses and mean Symptom Burden (SD shaded) over time for (**A**) BSGM Normal, showing post-meal power increases, and a sustained frequency band, (**B**) BSGM Delayed showing a delay in meal response until 2-3 hours post-meal, indicated by amplitude increase, and (**C**) Low Stability/Low Amplitude showing a lack of stable activity (illustrated by the yellow scatter) with minimal amplitude change over time. The Sankey Plot (**D**) illustrates the lack of relationship between Functional Dyspepsia and Gastroparesis patients to BSGM Phenotypes.

There was no relationship between the clinical diagnoses of FD and gastroparesis and the physiological phenotypes revealed by BSGM, as demonstrated by the Sankey Plot in Figure 3D.

## Discussion

This study applied a new BSGM technique (Gastric Alimetry) in an adolescent population in order to determine whether meaningful new pediatric disease groupings could be delineated in FD and gastroparesis. The results demonstrated that FD and gastroparesis patients could not be separated by their symptoms, quality of life, functional disability scores, health psychology metrics, or BSGM metrics. In contrast, three distinct BSGM phenotypes emerged from the spectral analysis, showing meaningful clinical differences across most of these domains. The clinical diagnosis of FD or gastroparesis showed no relationship to the three BSGM phenotypes, indicating that they potentially constitute novel disease categories.

Our findings, supported by other research, found substantial overlap between FD and gastroparesis regarding symptom paterns, quality of life, and mental wellbeing.^10, 23^ The overlap between FD and gastroparesis poses difficulties to personalised care, particularly for predicting patient responses to management options. Consequently, there is an element of trial and error to current clinical decisions and patient care pathways,^24, 25^ which appears unavoidable without improved diagnostic tools to provide improved clinical biomarkers.

BSGM introduces objective phenotyping and symptom profiling and thus offers an alternate approach or a secondary diagnostic layer to GET.^9, 12, 26^

A known weakness of GET is that it does not dependably capture neuromuscular pathologies, such as injury to the interstitial cell of Cajal (ICC) networks, which can lead to dysrythmic gastric myoelectrical activity.^27^ A primary motivation for developing BSGM was to resolve this gap by introducing a test more specific for neuromuscular disorders, considered to be characterized here by the Low GA-RI / Low amplitude phenotype.^12, 28^ In this study on adolescents, this phenotype was identified in both gastroparesis and FD subjects and was notable for its more severe symptoms (particularly nausea), poorer health psychology, physical health and lower quality of life. These factors indicate that this phenotype likely distinguishes a more severely affected subset of pediatric patients requiring more intensive management approaches.

Patients with high symptom burden, unrelated to gastric amplitude, plausibly indicate a gut-brain axis relationship (*BSGM Normal* phenotype).^29^ In adults, research has found that patients in this category often experience higher psychological comorbidities;^24, 30–32^ however, in our adolescent study, anxiety and depression scores in this group were similar to the population average, indicating that other factors should also be considered.^33,34^ Patients with the *BSGM Delayed* group constitute a new category, with a temporal disconnect between the meal challenge and subsequent meal response (amplitude increase), with symptoms occurring predominantly during the lag phase. This group may indicate an underlying disordered accommodation disorder, of which a lag in emptying is a feature, or could be demonstrating generalised transient gastric hypomotility.^35–37^ The finding that seven patients diagnosed with gastroparesis had a normal spectral outcome (*BSGM Normal* phenotype) is significant and could indicate a focus on alternative pathophysiologies, including pyloric resistance.^38^

This study also included the recruitment of the first cohort of adolescent healthy control subjects. This data was applied to compare adolescent and adult control data from a separate study,^20^ demonstrating that adult reference intervals can provide an acceptable provisional BSGM interpretation framework. While this qualifying step enabled the delineation of the first pediatric spectral groupings, in future we recommend that age-specific pediatric reference intervals be prioritised. This need is supported by a modestly reduced GA-RI metric and visibly shorter meal response duration evident in adolescent controls in this first study, which may be accentuated when smaller meals are employed in pediatric populations.^39^

Several limitations are acknowledged. While our sample size was sufficient for delineating novel pediatric disorder subgroups, further studies with larger patient groups are now desirable to confirm and expand the outcomes reported here. This study focused on FD confirmed by Rome IV, and a normal gastric emptying scintigraphy, and gastroparesis patients only. Given the prevalence of common paediatric DGBI conditions such as functional abdominal pain, chronic nausea and vomiting,^1^ expanding BSGM research into more conditions is important. In addition, studies linking the newly defined subgroups to therapeutic outcomes comprise a critical next step to further defining their clinical importance. The limited male representation in the patient group is unsurprising as the prevalence of these disorders are higher in females.^40–42^

In conclusion, this first study of BSGM in a pediatric population has identified novel subgroups in adolescents with chronic gastroduodenal symptoms. These groups showed meaningful clinical differences, which were absent when comparing FD and gastroparesis. These findings indicate that BSGM can provide valuable data to classify disease phenotypes within these complex conditions, which could support differential treatment approaches, follow-ups, and monitoring.

## Data Availability

Data sharing and data use are governed by the New Zealand Health and Disability Ethics Committee. The majority of the data is available in the manuscript however all requests for additional data can be made to the corresponding author. Requests will be granted if the proposed use aligns with the ethical approval for the study and relevant ethical approvals have been obtained, including approval from a New Zealand Ethical Committee and Childrens Hospital of Philadelphia IRB Pennsylvania USA.  

---

**Figure S1.**
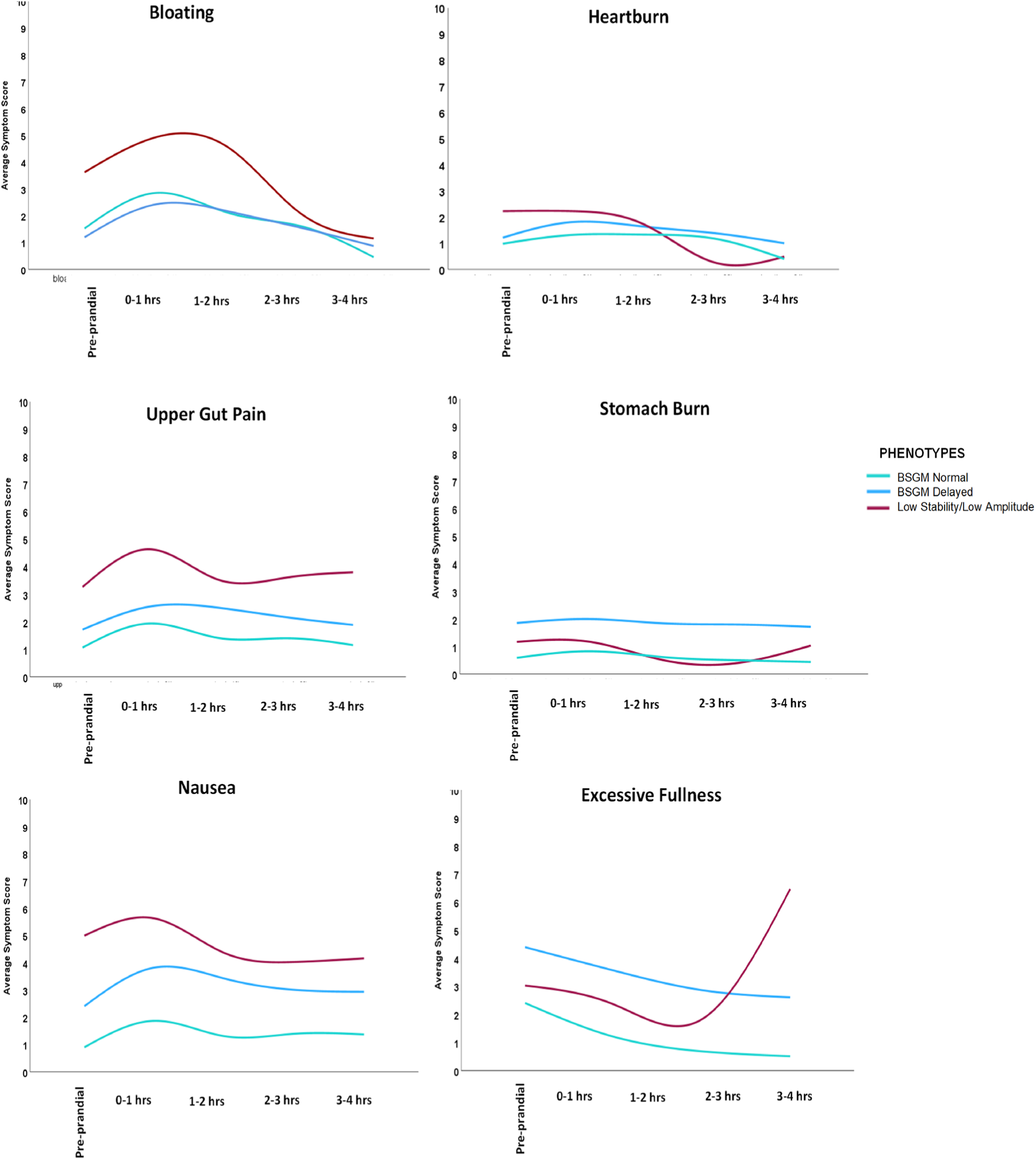
BSGM phenotypes showing within-test individual symptom curves

**Table S1.** Pediatric Healthy Control and Patient Demographics.

| VARIABLES | Healthy Controls<br>(N=31) | Patients<br>(n=25) | *P*value |
| --- | --- | --- | --- |
| **Demographics** |  |  |  |
| Sex (M (%)) | 19 (61) | 1 (4) | <.001 |
| Age years (median (IQR)) | 14.7 (13-16) | 16.1 (14-18) | 0.02 |
| Ethnicity (N (%)) |  |  |  |
| White/European | 22 (71) | 20 (80) | - |
| Māori<sup>a</sup> | 1 (3) | - | - |
| Pacific/Hawaiian | 2 (6.5) | - | - |
| Black/African American | 2 (6.5) | 2 (8) | - |
| Hispanic/Latinx | - | 3 (12) | - |
| Chinese | 2 (6.5) | - | - |
| Other | 2 (6.5) | - | - |
| BMI Kg/m<sup>2</sup> (mean (SD)) | 21.1 (3.28) | 21.2 (2.63) | 0.2 |
| **Diagnosis** |  |  |  |
| Gastroparesis | - | 15 | - |
| Functional Dyspepsia | - | 10 | - |
| EPS<sup>b</sup> | - | 5 | - |
| PDS<sup>c</sup> | - | 5 | - |
| **Quality** |  |  |  |
| % of Meal Completion (median (IQR)) | 100 | 66.7 (40-100) | <.001 |
| % Artifact (mean (SD)) | 24.4 (12.7) | 23.9 (12.91) | 0.6 |
| Impedance (median (IQR)) | 108.9 (67.4-124.2) | 131.2 (91.3-178.7) | 0.04 |

<sup>a</sup>Māori: Indigenous population of New Zealand; <sup>b</sup>EPS: Epigastric Pain Syndrome, <sup>c</sup>PDS: Post Prandial Distress Syndrome

**Table S2.** Total Symptom Burden and Individual Symptom Scores, and Clinical Symptoms, Quality of Life, Functional Disability, and Mental Wellbeing Questionnaire Outcomes for Healthy Controls and Patients.

| VARIABLES |  | Healthy Controls | Patients | P value |
| --- | --- | --- | --- | --- |
| (median (IQR)) |  | N=31 | N=25 |  |
| Total Symptom Burden |  | 1.2 (0.0-3.6) | 16.4 (5.3-33.1) | <.001 |
| Individual Symptoms | Early Satiety | 1 (0.0-2.0) | 4 (1.0-6.0) | 0.04 |
|  | Excessively Full | 0 (0.0-.55) | 2.55 (0.18-5.7) | <.001 |
|  | Bloating | 0 | 1.8 (0.0-3.95) | <.001 |
|  | Heartburn | 0 | 0.3 (0.0-2.45) | <.001 |
|  | Nausea | 0 | 2.3 (0.15-6.5) | <.001 |
|  | Upper Gut Pain | 0 | 2.8 (0.03-5.2) | <.001 |
|  | Stomach Burn | 0 | 1.2 (0.0-4.3) | <.001 |
| **Patient Completed Questionnaires** |  |  |  |  |
| Gastrointestinal Symptoms | ↓ | 89.8 (83.4-95.6) | 53.72 (38.51-70.61) | 0.02 |
| Nausea Severity Scale | ↑ | 0.0 (0.0-14.0) | 20.4 (10.2-29.7) | 0.003 |
| Abdominal Pain Index | ↑ | 0 | 23.0 (14.2-31.0) | <.001 |
| Functional Disability Index | ↑ | 0.0 (0.0-1.0) | 20.0 (9.0-30.25) | <.001 |
| Quality of Life | ↓ | 97.0 (85.0-100) | 57.0 (42.0-71.0) | <.001 |
| Anxiety | ↑ | 38.0 (33.5-52.5) | 53.6 (43.2-59.5) | <.001 |
| Depression | ↑ | 43.8 (33.5-52.5) | 50.9 (43.5-59.5) | 0.32 |

↑↓: indicates the direction of severity for each questionnaire

**Table S3.** Comparison between Pediatric and Adult Controls for average BMI-<u>adjusted amplitude, PGF, GA-RI, f</u>f-AR and meal response ratio.

| BSGM Spectral Metrics | Pediatric Controls | Adult Controls | *P* value |
| --- | --- | --- | --- |
|  | N=31 | N=110 |  |
| Average BMI-Adj Amplitude (mean (SD)) | 34.58 (8.8) | 40.97 (17.4) | 0.12 |
| Average PGF<sup>a</sup> (mean (SD)) | 3.03 (.19) | 3.04 (.20) | 0.93 |
| Average GA-RI<sup>b</sup> (median (IQR)) | 0.35 (.22-.43) | 0.50 (.39-.64) | <.001 |
| Average ff-AR<sup>c</sup> (mean (SD)) | 2.24 (.78) | 1.96 (.89) | 0.02 |
| Meal Response Ratio (median (IQR)) | 1.44 (1.32-1.82) | 1.10 (.94-1.38) | <.001 |

<sup>a</sup>PGF: Principal Gastric Frequency; <sup>b</sup>GA-RI: Gastric Alimetry Rhythm Index; <sup>c</sup>ff-AR: fed to fasted Amplitude Ratio

**Table S4.** Demographics and Test Quality for Patient Phenotypes.

**eTable 4.** Demographics and Test Quality for Patient Phenotypes
| Variables | BSGM Normal<br>(N=10) | BSGM Delayed<br>(N=8) | Low Stability/<br>Low Amplitude<br>(N=7) | *P* value |
| --- | --- | --- | --- | --- |
| Sex (M (%)) | 0 (0) | 1 (12) | 0 (0) | - |
| Age years (median (IQR)) | 16.5 (12.7-19) | 17 (16.25-18) | 16 (14-18) | 0.63 |
| BMI kg/m<sup>2</sup> (mean (SD)) | 20.4 (1.3) | 22.1 (2.9) | 21.8 (2.0) | 0.4 |
| % Meal Complete (median (IQR)) | 95 (65-100) | 65 (42.5-100) | 50 (40-90) | 0.59 |
| % Artifact (mean (SD)) | 21.66 (11.1) | 21.45 (13.7) | 25.02 (12.3) | 0.36 |
| Impedance (median (IQR)) | 116.95 (86.5-146.0) | 105.25 (57.9-181.5) | 136.7 (107.1-230.0) | 0.26 |

## REFERENCES

1. Boronat AC, Ferreira-Maia AP, Matijasevich A, et al. Epidemiology of functional gastrointestinal disorders in children and adolescents: A systematic review. World J Gastroenterol 2017;23:3915–3927.

2. Dipasquale V, Corica D, Gramaglia SMC, et al. Gastrointestinal symptoms in children: Primary care and specialist interface. International journal of clinical practice 2018;72:e13093.

3. Holtman GA. Diagnostic strategies in children with chronic gastrointestinal symptoms in primary care. Volume PhD: Rijksuniversiteit Groningen, 2016.

4. 4. Drossman D, Chang L, Chey W, et al. Rome IV Pediatric Functional Gastrointestinal Disorders – Disorders of Gut-Brain Interaction, First Edition Raleigh, North Carolina: Rome Foundation, 2016.

5. Friesen CA, Rosen JM, Schurman JV. Prevalence of overlap syndromes and symptoms in pediatric functional dyspepsia. BMC Gastroenterol 2016;16:75.

6. Talley NJ, Ford AC. Functional Dyspepsia. N Engl J Med 2015;373:1853–63.

7. Wong GK, Shulman RJ, Chumpitazi BP. Gastric emptying scintigraphy results in children are affected by age, anthropometric factors, and study duration. Neurogastroenterology and Motility: the official journal of the European Gastrointestinal Motility Society 2015;27:356–362.

8. Kim BJ, Kuo B. Gastroparesis and Functional Dyspepsia: A Blurring Distinction of Pathophysiology and Treatment. J Neurogastroenterol Motil 2019;25:27–35.

9. Wang WJ, Foong D, Calder S, et al. Gastric Alimetry improves patient phenotyping in gastroduodenal disorders compared to gastric emptying scintigraphy alone. medRxiv 2023:2023.05.18.23290134.

10. Karamanolis G, Caenepeel P, Arts J, et al. Determinants of symptom patern in idiopathic severely delayed gastric emptying: gastric emptying rate or proximal stomach dysfunction? Gut 2007;56:29–36.

11. Tack J, Carbone F. Functional dyspepsia and gastroparesis. Curr Opin Gastroenterol 2017;33:446–454.

12. Gharibans AA, Calder S, Varghese C, et al. Gastric dysfunction in patients with chronic nausea and vomiting syndromes defined by a noninvasive gastric mapping device. Science Translational Medicine 2022;14:eabq3544.

13. Wang W, Foong D, Calder S, et al. Gastric Alimetry Expands Patient Phenotyping in Gastroduodenal Disorders Compared with Gastric Emptying Scintigraphy. ACG 2023; October 1-11.

14. Xu W, Williams L, Serebatnam G, et al. Gastric Alimetry testing and healthcare economic analysis in nausea and vomiting syndromes. medRxiv 2023:2023.09.07.23295185.

15. Humphrey G, Schamberg G, Keane C, et al. Towards age-specific normative values for body surface gastric mapping in pediatrics Gastroenterology 2023;164.

16. Humphrey G, Schamberg G, Keane C, et al. Translation of body surface gastric mapping to pediatric populations: Feasibility and initial results. Gastroenterology (New York, N.Y. 1943) 2023;164:S–30.

17. Cuschieri S. The STROBE guidelines. Saudi J Anaesth 2019;13:S31–S34.

18. 18. Reddivari A, Mehta P. Gastroparesis. National Library of Medicine, 2022; https://www.ncbi.nlm.nih.gov/books/NBK551528/

19. Abell TL, Camilleri M, Donohoe K, et al. Consensus recommendations for gastric emptying scintigraphy: a joint report of the American Neurogastroenterology and Motility Society and the Society of Nuclear Medicine. J Nucl Med Technol 2008;36:44–54.

20. Varghese C, Schamberg G, Calder S, et al. Normative values for body surface gastric mapping evaluations of gastric motility using Gastric Alimetry: spectral analysis. American Journal of Gastroenterology 2023;118:1047–1057.

21. Sebaratnam G, Karulkar N, Calder S, et al. Standardized system and App for continuous patient symptom logging in gastroduodenal disorders: Design, implementation, and validation. Neurogastroenterology & Motility 2022;34:e14331.

22. Schamberg G, Varghese C, Calder S, et al. Revised spectral metrics for body surface measurements of gastric electrophysiology. Neurogastroenterology & Motility 2023;35:e14491.

23. Mandarino FV, Testoni SGG, Barchi A, et al. Imaging in Gastroparesis: Exploring Innovative Diagnostic Approaches, Symptoms, and Treatment. Life (Basel, Switzerland) 2023;13:1743.

24. Stanghellini V, Chan FKL, Hasler WL, et al. Gastroduodenal Disorders. Gastroenterology 2016;150:1380–1392.

25. Sebaratnam G, Law M, Broadbent E, et al. “It’s a helluva journey”: A qualitative study of patient and clinician experiences of nausea and vomiting syndromes and functional dyspepsia. Frontiers in Psychology 2023;14.

26. Xu W, Gharibans AA, Calder S, et al. Defining and Phenotyping Gastric Abnormalities in Long-Term Type 1 Diabetes Using a Novel Body Surface Gastric Mapping Device. Gastro Hep Advances 2023;2:1120–1132.

27. Pasricha PJ, Grover M, Yates KP, et al. Functional Dyspepsia and Gastroparesis in Tertiary Care are Interchangeable Syndromes With Common Clinical and Pathologic Features. Gastroenterology 2021;160:2006–2017.

28. Lin Z, Sarosiek I, Forster J, et al. Association of the status of interstitial cells of Cajal and electrogastrogram parameters, gastric emptying and symptoms in patients with gastroparesis. Neurogastroenterology and motility 2010;22:56–e10.

29. Koloski NA, Jones M, Kalantar J, et al. The brain–gut pathway in functional gastrointestinal disorders is bidirectional: a 12-year prospective population-based study. Gut 2012;61:1284–1290.

30. Tarbell SE, Shaltout HA, Wagoner AL, et al. Relationship among nausea, anxiety, and orthostatic symptoms in pediatric patients with chronic unexplained nausea. Experimental brain research 2014;232:2645–2650.

31. Waters AM, Waters AM, Schilpzand E, et al. Functional Gastrointestinal Symptoms in Children with Anxiety Disorders. Journal of abnormal child psychology 2013;41:151–163.

32. Schutyser W, Cruyt L, Vulsteke J-B, et al. The role of high-resolution manometry in the assessment of upper gastrointestinal involvement in systemic sclerosis: a systematic review. Clinical Rheumatology 2020;39:149–157.

33. Health Measures. Available PROMIS Measures for Pediatric Self Report. Volume 2023, 2023.

34. Wu JC. Community-based study on psychological comorbidity in functional gastrointestinal disorder. Journal of Gastroenterology and Hepatology 2011;26:23–26.

35. O’Grady G, Carbone F, Tack J. Gastric sensorimotor function and its clinical measurement. Neurogastroenterology & Motility 2022:e14489.

36. O’Grady G, Varghese C, Schamberg G, et al. An Initial Phenotype Set for the Assessment of Gastroduodenal Disorders with Gastric Alimetry®, 2023.

37. Febo-Rodriguez L, Chumpitazi BP, Sher AC, et al. Gastric accommodation: Physiology, diagnostic modalities, clinical relevance, and therapies. Neurogastroenterology & Motility 2021;33:e14213.

38. Wellington J, Scot B, Kundu S, et al. Effect Effect of endoscopic pyloric therapies for patients with nausea and vomiting and functional obstructive gastroparesis. Autonomic neuroscience 2017;202:56–61.

39. Huang I-H, Calder S, Gharibans AA, et al. Meal Effects on Gastric Bioelectrical Activity Utilizing Body Surface Gastric Mapping in Healthy Subjects. medRxiv 2023:2023.10.31.23296947.

40. Spee LAA, Lisman-Van Leeuwen Y, Benninga MA, et al. Prevalence, characteristics, and management of childhood functional abdominal pain in general practice. Scandinavian Journal of Primary Health Care 2013;31:197–202.

41. Sperber AD, Bangdiwala SI, Drossman DA, et al. Worldwide Prevalence and Burden of Functional Gastrointestinal Disorders, Results of Rome Foundation Global Study. Gastroenterology 2020.

42. Clara de Bruijn, Anne Geijtenbeek, Pamela D. Browne, et al. Children with functional gastrointestinal disorders with and without co-existing nausea: A comparison of clinical and psychological characteristics. Neurogastroenterology & Motility 2023:e14591.

